# Associations with covid-19 hospitalisation amongst 406,793 adults: the UK Biobank prospective cohort study

**DOI:** 10.1101/2020.05.06.20092957

**Authors:** Anthony P Khawaja, Alasdair N Warwick, Pirro G Hysi, Alan Kastner, Andrew Dick, Peng T Khaw, Adnan Tufail, Paul J Foster, Kay-Tee Khaw

## Abstract

**OBJECTIVES:** To identify the sociodemographic, lifestyle, comorbidity and antihypertensive medication associations with the development of hospitalisation with covid-19 in an English population.

**DESIGN:** Prospective cohort study

**SETTING:** The population-based UK Biobank study was linked to English covid-19 test results.

**PARTICIPANTS:** Individuals resident in England and alive in 2020.

**MAIN OUTCOME MEASURES:** Cases (n=605) were defined by a positive covid-19 test result conducted between 16^th^ March and 16^th^ April 2020, during a restricted testing policy for hospitalised individuals with severe disease.

**RESULTS:** A total of 406,793 participants were included. Mean age on 1^st^ January 2020 was 68 years (range 48 to 85 years). 55% were women. In multivariable models, major independent risk factors for hospitalisation with covid-19 were male sex (odds ratio 1.52; 95% confidence interval 1.28 to 1.81; P<0.001), South Asian ethnicity (2.02; 1.28 to 3.17; P=0.002) or black ethnicity (3.09; 2.18 to 4.38; P<0.001) compared to white ethnicity, greater residential deprivation (1.92 for most deprived quartile compared to least deprived quartile; 1.50 to 2.47; P<0.001), higher BMI (2.04 for BMI >35 compared to <25 Kg/m^2^; 1.50 to 2.77; P<0.001), former smoking (1.39 compared to never smoked; 1.16 to 1.66; P<0.001), and comorbidities hypertension (1.28; 1.06 to 1.53; P=0.009) and chronic obstructive pulmonary disease (1.81; 1.34 to 2.44; P<0.001). Increased risk was observed with increasing number of antihypertensive medications used rather than any individual class.

**CONCLUSION:** Understanding why these factors confer increased risk of severe covid-19 in the population may help elucidate the underlying mechanisms as well as inform strategy and policy to prevent this disease and its consequences. We found no evidence of increased risk with specific classes of antihypertensive medication.

## Introduction

The UK has been one of the most severely affected countries in the coronavirus disease 2019 (covid-19) pandemic. As of 5^th^ May 2020, the total number of lab-confirmed UK cases was 194,990 and there have been 29,427 covid-19 associated deaths.^1^ Examination of the characteristics of people hospitalised with covid-19 and their outcomes has given some insight into the epidemiology of the disease.^2–4^ Reports have suggested that ethnicity^3,5–9^ and residential deprivation^10^ are associated with death from covid-19, prompting a rapid review to be led by Public Health England.^11^ There is also interest in whether use of antihypertensive medications that affect the renin-angiotensin system can increase the risk of severe disease.^12^ However, hospital-based studies lack a population-based control group and therefore cannot accurately determine risk factors for onset of severe disease, or easily test for independence of risk factors. It remains uncertain, for example, whether ethnicity is associated with the incidence of severe covid-19 or with a worse outcome after developing severe disease.^13^ Understanding population-level risk factors for covid-19 is of major interest to the general public and public health bodies, and will underpin strategies for population-wide disease prevention.

The UK Biobank study has been following half a million participants for over a decade and has collected rich exposure data during this time. Recently, covid-19 test results for hospitalised patients have been made available by Public Health England. We present an analysis identifying the sociodemographic, lifestyle and comorbidity factors that are associated with the development of severe covid-19 in the English population. Additionally, we examine whether antihypertensive medication use is associated with risk of severe covid-19.

## Methods

### UK Biobank Assessment

The UK Biobank is a very large multisite cohort study that recruited over half a million UK residents aged 40 to 69 years for a baseline examination between 2006 and 2010. Detailed study protocols are available online (http://www.ukbiobank.ac.uk/resources/). Follow-up examinations were carried out in subsamples (fig 1).

**Figure 1:**
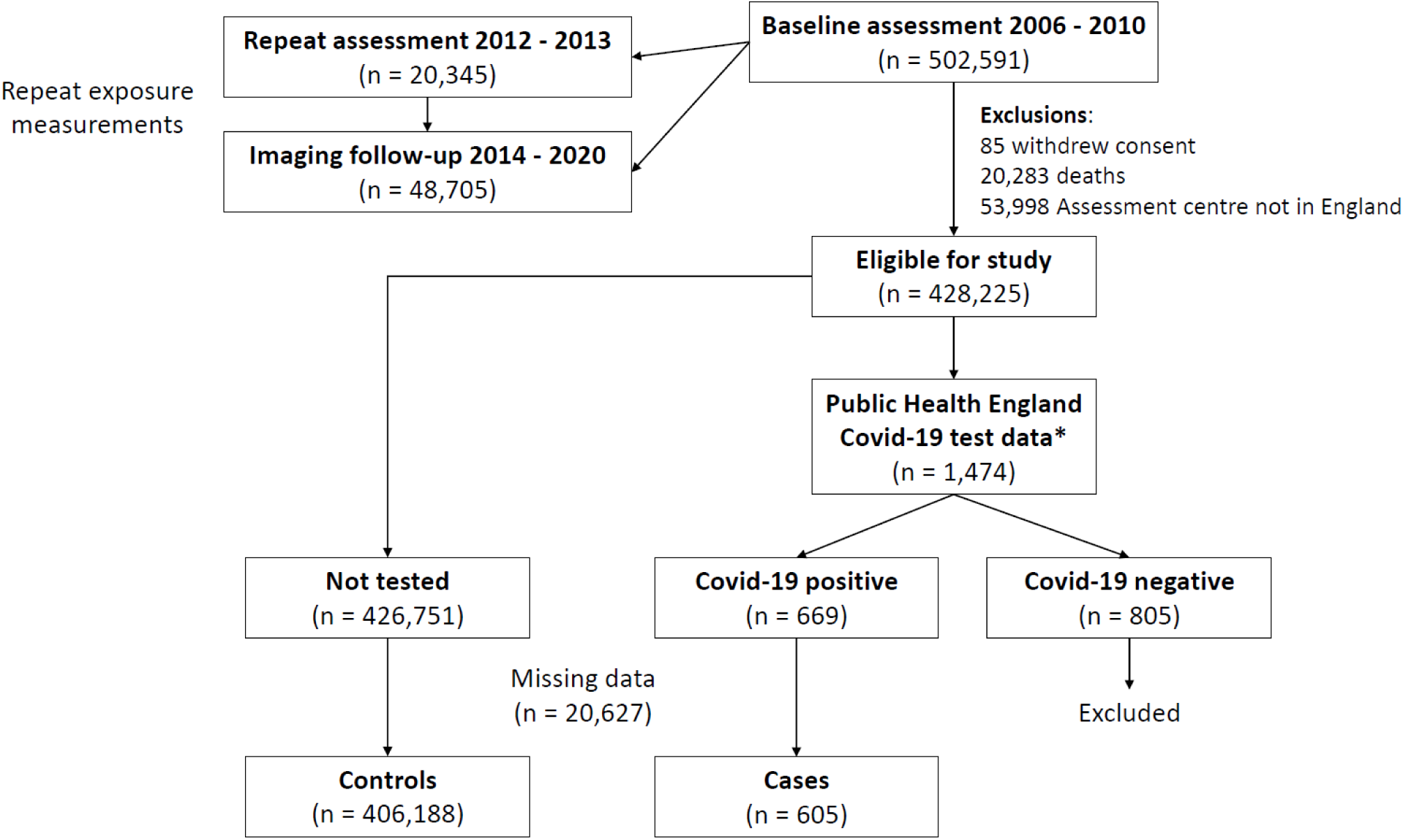
Chart presenting participant flow and ascertainment of case and control status. * Test data were from 16^th^ March 2020 onwards, when a restricted testing policy was in place; testing was reserved for hospitalised individuals with assumed severe covid-19.

Participants completed a touch-screen self-administered questionnaire, an interview and physical measurements (supplementary table S1). Common cardiorespiratory comorbidity history was ascertained using a combination of self-report (supplementary table S1) and linked hospital episode statistic (HES) data. Participants were considered to have a history of each comorbidity if there was either evidence of self-report or from HES data, or both. All medication use was ascertained by interview and each drug allocated a code. Please see the supplementary materials (UK Biobank Medication Codes) for a list of codes we used for each antihypertensive medication class. For participants with medication data at follow-up visits, we considered the status at the most recent interview.

Patients or the public were not involved in the design, or conduct, or reporting, or dissemination plans of our research. There are no specific plans to disseminate the results to study participants or patient organisations.

### Covid-19 Status Ascertainment

Hospitalisation with covid-19 was ascertained via record linkage to covid-19 test results carried out in England, as provided to UK Biobank by Public Health England. Detailed information on the testing and linkage are available.^14^ Linkage was carried out to test results from 16^th^ March 2020 onwards, as testing was largely restricted to hospitalised patients after this date. We are considering a positive result as a surrogate for severe covid-19 disease which required hospitalisation. For participants with multiple covid-19 tests, cases were defined as all participants with at least one positive test. Assuming severe covid-19 to be relatively uncommon, we considered the remainder of the cohort who had not been tested to be controls. We excluded participants that were tested but without a positive covid-19 test in case a proportion were false negatives and given the abundance of controls already available. Data were also available for whether hospitalisation was documented on the test request form. Absence of hospitalisation documentation on the request form was not considered indicative of lack of hospitalisation given the testing policy. However, we additionally conducted a sensitivity analysis excluding cases without hospitalisation indicated on the test request form.

### Statistical Analysis

Participants who died before 2020 or did not attend an assessment centre in England were excluded given they could not become cases. We analysed all participants with available data and no sample size calculation was carried out. We compared sociodemographic, comorbidity and antihypertensive medication data between cases and controls using univariable logistic regression. We further examined antihypertensive medications together in a multivariable logistic regression model, both with and without hypertension diagnosis as a covariable. Similarly, we examined all comorbidities together in a multivariable model. Factors which were nominally significant were then all assessed together in a single multivariable model, and those with strong associations, together with age, were taken forward to the final parsimonious model. As a sensitivity analysis, we re-ran the final model following exclusion of cases without documentation of hospitalisation on the specimen request form. We did not adjust confidence intervals or P-values for multiple testing. All analyses were conducted with Stata software, version 15.1 (StataCorp).

## Results

Figure 1 presents participant flow. In total, 406,793 participants were included in this analysis (605 cases and 406,188 controls). Mean age at recruitment was 56 years (standard deviation 8; range 38 to 73 years) and mean age on 1^st^ January 2020 was 68 years (8; 48 to 85 years). 55% were women. Table 1 presents baseline data for the cohort. Cases were significantly older, more likely to be men and more likely to be Black or South Asian. Cases had lower educational attainment and resided in more deprived locations; 40% of cases were in the most deprived quartile of Townsend deprivation index. Cases weighed more, had a higher body mass index (BMI) and a slightly higher diastolic blood pressure. Cases consumed alcohol less frequently and were more likely to be former smokers. All comorbidities were more frequent in cases. Nearly half of cases had a history of hypertension. Cases were more likely to use all classes of antihypertensive medication. Univariable regression results for all examined factors are presented in supplementary table S2.

**Table 1:**
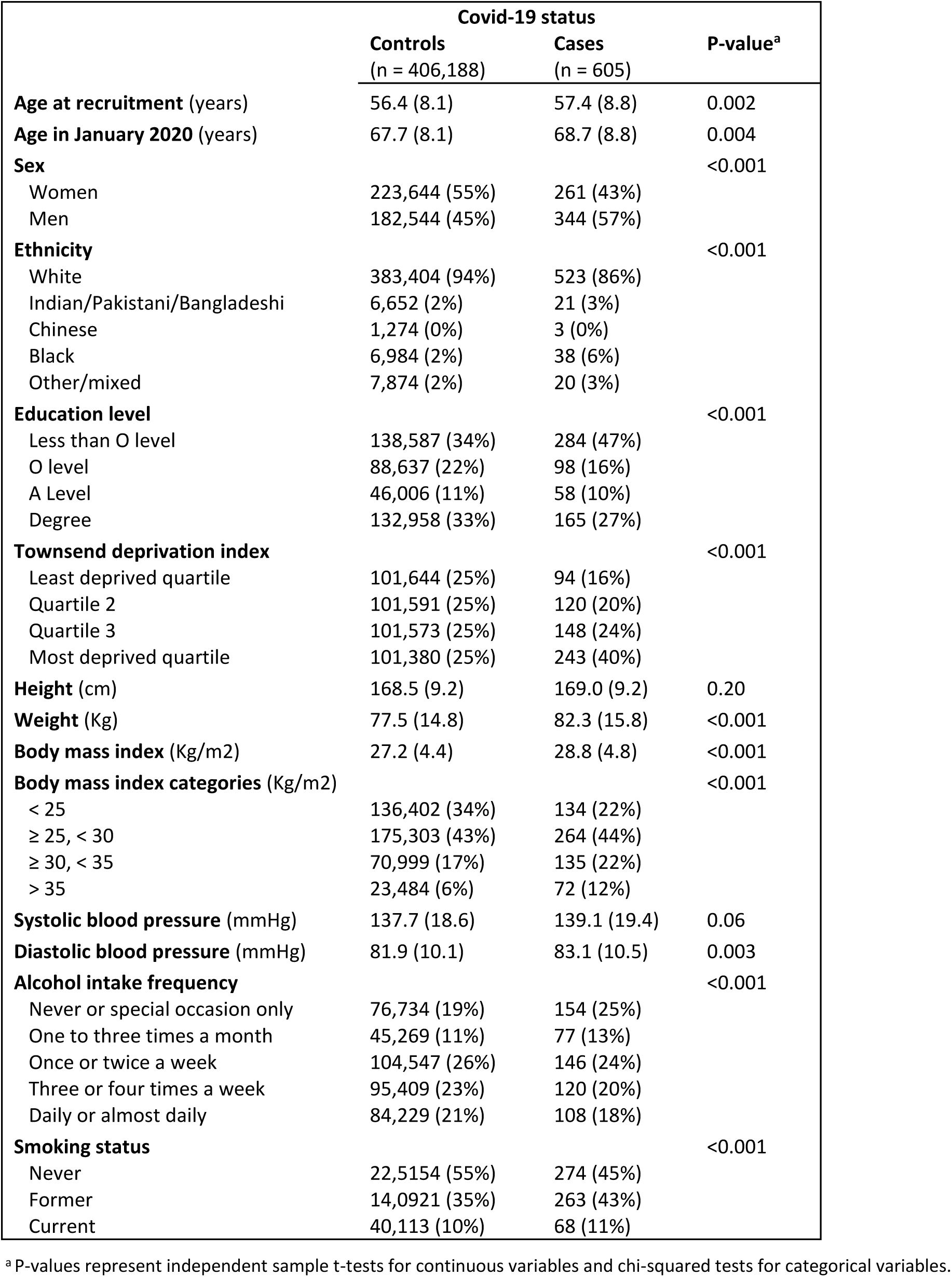
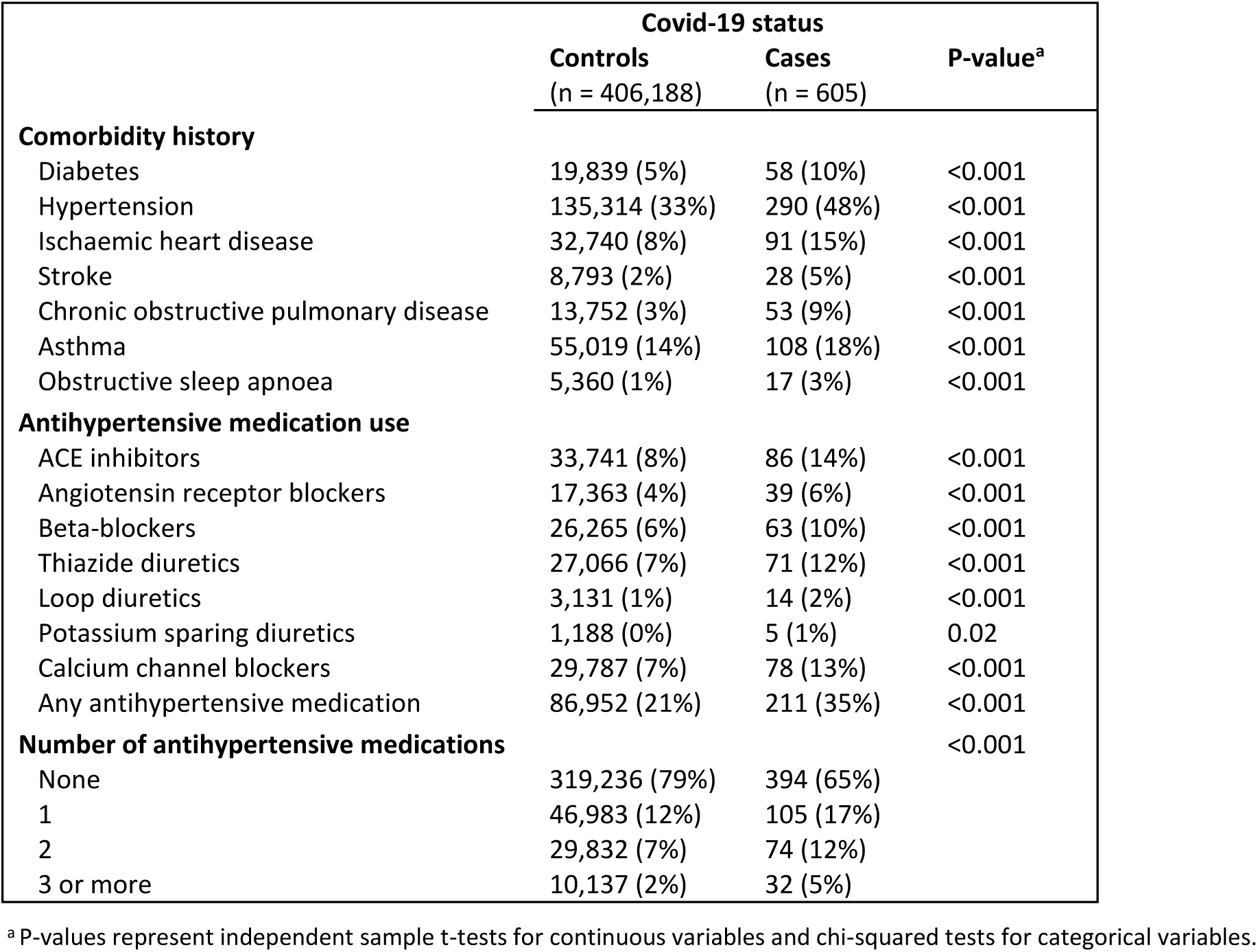
Sociodemographic factors, comorbidity history and antihypertensive medication use in cases and controls.

In a multivariable model with all antihypertensive classes together, adjusted for age, sex and ethnicity, the strength of all medication associations were markedly reduced (table 2). After further adjusting for hypertension, loop diuretics were the only medication class significant at the 5% level, but hypertension was strongly associated with an increased risk (table 2; odds ratio 1.41, 95% confidence interval 1.15 to 1.72; P=0.001). When considering the number of antihypertensive medications and hypertension as a comorbidity together, both terms were significantly associated with an increased risk (table 2). Participants using 1, 2 or 3+ antihypertensive medications were at 37%, 44% and 75% increased odds of covid-19 hospitalisation compared to participants not using antihypertensives. When all comorbidities were examined together in a single multivariable model, hypertension (1.44; 1.22 to 1.72; P<0.001) and chronic obstructive pulmonary disease (COPD; 2.10, 1.55 to 2.84; P<0.001) were highly significantly associated, and borderline associations observed for ischemic heart disease and stroke (supplementary table S3).

**Table 2:** Results from 3 multivariable regression models presenting associations between antihypertensive medication classes and hospitalisation with covid-19 (n = 406,793).

|  | Model 1<sup>a</sup> |  | Model 2<sup>b</sup> |  | Model 3<sup>c</sup> |  |
| --- | --- | --- | --- | --- | --- | --- |
|  | OR (95% CI) | P-value | OR (95% CI) | P-value | OR (95% CI) | P-value |
| **ACE inhibitors** | 1.34 (1.04 to 1.73) | 0.02 | 1.17 (0.90 to 1.52) | 0.23 | - | - |
| **Angiotensin receptor blockers** | 1.13 (0.80 to 1.60) | 0.48 | 1.00 (0.70 to 1.42) | 0.99 | - | - |
| **Beta-blockers** | 1.20 (0.91 to 1.59) | 0.19 | 1.13 (0.85 to 1.49) | 0.41 | - | - |
| **Thiazide diuretics** | 1.44 (1.10 to 1.90) | 0.008 | 1.30 (0.99 to 1.72) | 0.06 | - | - |
| **Loop diuretics** | 2.16 (1.23 to 3.82) | 0.008 | 2.11 (1.20 to 3.71) | 0.01 | - | - |
| **Potassium sparing diuretics** | 1.55 (0.62 to 3.87) | 0.35 | 1.54 (0.62 to 3.85) | 0.36 | - | - |
| **Calcium channel blockers** | 1.23 (0.95 to 1.59) | 0.12 | 1.10 (0.85 to 1.44) | 0.46 | - | - |
| **Hypertension comorbidity** | - | - | 1.41 (1.15 to 1.72) | 0.001 | 1.33 (1.07 to 1.65) | 0.011 |
| **Number of antihypertensive medications** | - | - | - | - |  |  |
| None |  |  |  |  | Ref |  |
| 1 |  |  |  |  | 1.37 (1.05 to 1.78) | 0.020 |
| 2 |  |  |  |  | 1.44 (1.07 to 1.94) | 0.02 |
| 3 or more |  |  |  |  | 1.75 (1.17 to 2.61) | 0.006 |
| *Test for trend* |  |  |  |  |  | 0.001 |

Abbreviations: CI = confidence interval; OR = odds ratio.
<sup>a</sup> Model 1: All antihypertensive medication classes, adjusted for age, sex and ethnicity.
<sup>b</sup> Model 2: All antihypertensive medication classes plus hypertension comorbidity status, adjusted for age, sex and ethnicity.
<sup>c</sup> Model 3: Number of antihypertensive medications and hypertension comorbidity status, adjusted for age, sex and ethnicity.

We then examined all variables showing significant association in the above analysis together in a single multivariable model (table 3). The strongest associations with severe covid-19 were male sex (1.52, 1.28 to 1.81; P<0.001), South Asian ethnicity (2.02, 1.28 to 3.17; P=0.002) or black ethnicity (3.09, 2.18 to 4.38; P<0.001) ethnicity (compared to White ethnicity), greater residential deprivation (1.92 for most deprived quartile compared to least deprived quartile, 1.50 to 2.47; P<0.001), higher BMI compared to <25 Kg/m^2^: 1.26 for ≥25-<30, 1.37 for ≥30-<35 and 2.04 for BMI >35 Kg/m^2^, former compared to never smoking (1.39, 1.16 to 1.66; P<0.001), and comorbidities hypertension (1.28, 1.06 to 1.53; P=0.009) and COPD (1.81, 1.34 to 2.44; P<0.001). All factors with strong association, together with age, were taken forward to a final parsimonious multivariable model (supplementary table S4) and visualized in figure 2. A sensitivity analysis excluding cases without clear evidence of hospitalisation on the specimen request form (n=90) gave similar results to the parsimonious model (supplementary table S5).

**Table 3:**
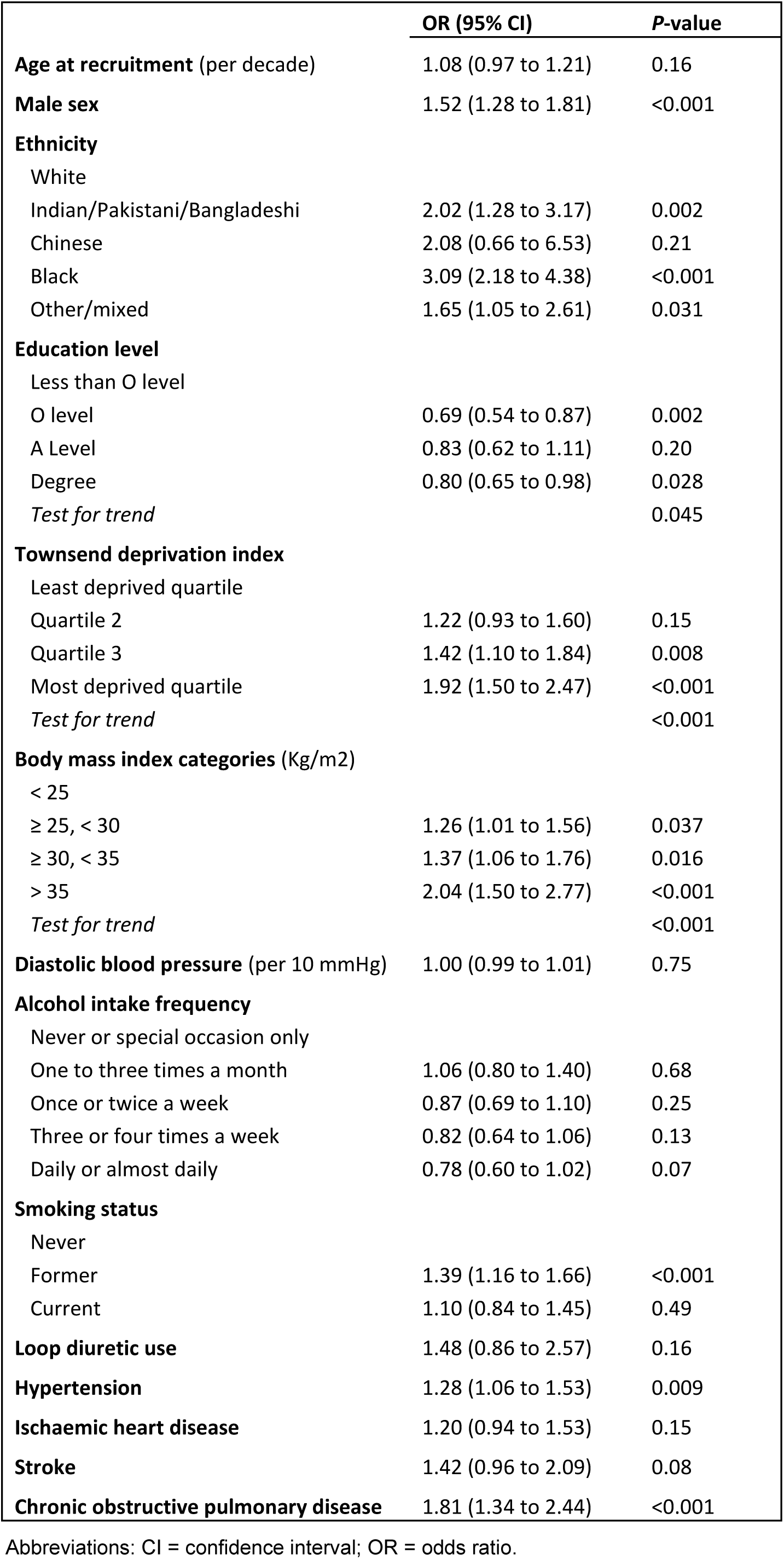
Results for a multivariable model examining all presented variables together for association with hospitalisation with covid-19 (n = 406,793).

|  | OR (95% CI) | P-value |
| --- | --- | --- |
| **Age at recruitment** (per decade) | 1.08 (0.97 to 1.21) | 0.16 |
| **Male sex** | 1.52 (1.28 to 1.81) | <0.001 |
| **Ethnicity** |  |  |
| White |  |  |
| Indian/Pakistani/Bangladeshi | 2.02 (1.28 to 3.17) | 0.002 |
| Chinese | 2.08 (0.66 to 6.53) | 0.21 |
| Black | 3.09 (2.18 to 4.38) | <0.001 |
| Other/mixed | 1.65 (1.05 to 2.61) | 0.031 |
| **Education level** |  |  |
| Less than O level |  |  |
| O level | 0.69 (0.54 to 0.87) | 0.002 |
| A Level | 0.83 (0.62 to 1.11) | 0.20 |
| Degree | 0.80 (0.65 to 0.98) | 0.028 |
| *Test for trend* |  | 0.045 |
| **Townsend deprivation index** |  |  |
| Least deprived quartile |  |  |
| Quartile 2 | 1.22 (0.93 to 1.60) | 0.15 |
| Quartile 3 | 1.42 (1.10 to 1.84) | 0.008 |
| Most deprived quartile | 1.92 (1.50 to 2.47) | <0.001 |
| *Test for trend* |  | <0.001 |
| **Body mass index categories** (Kg/m<sup>2</sup>) |  |  |
| < 25 |  |  |
| ≥ 25, < 30 | 1.26 (1.01 to 1.56) | 0.037 |
| ≥ 30, < 35 | 1.37 (1.06 to 1.76) | 0.016 |
| > 35 | 2.04 (1.50 to 2.77) | <0.001 |
| *Test for trend* |  | <0.001 |
| **Diastolic blood pressure** (per 10 mmHg) | 1.00 (0.99 to 1.01) | 0.75 |
| **Alcohol intake frequency** |  |  |
| Never or special occasion only |  |  |
| One to three times a month | 1.06 (0.80 to 1.40) | 0.68 |
| Once or twice a week | 0.87 (0.69 to 1.10) | 0.25 |
| Three or four times a week | 0.82 (0.64 to 1.06) | 0.13 |
| Daily or almost daily | 0.78 (0.60 to 1.02) | 0.07 |
| **Smoking status** |  |  |
| Never |  |  |
| Former | 1.39 (1.16 to 1.66) | <0.001 |
| Current | 1.10 (0.84 to 1.45) | 0.49 |
| **Loop diuretic use** | 1.48 (0.86 to 2.57) | 0.16 |
| **Hypertension** | 1.28 (1.06 to 1.53) | 0.009 |
| **Ischaemic heart disease** | 1.20 (0.94 to 1.53) | 0.15 |
| **Stroke** | 1.42 (0.96 to 2.09) | 0.08 |
| **Chronic obstructive pulmonary disease** | 1.81 (1.34 to 2.44) | <0.001 |

Abbreviations: CI = confidence interval; OR = odds ratio.

**Figure 2:**
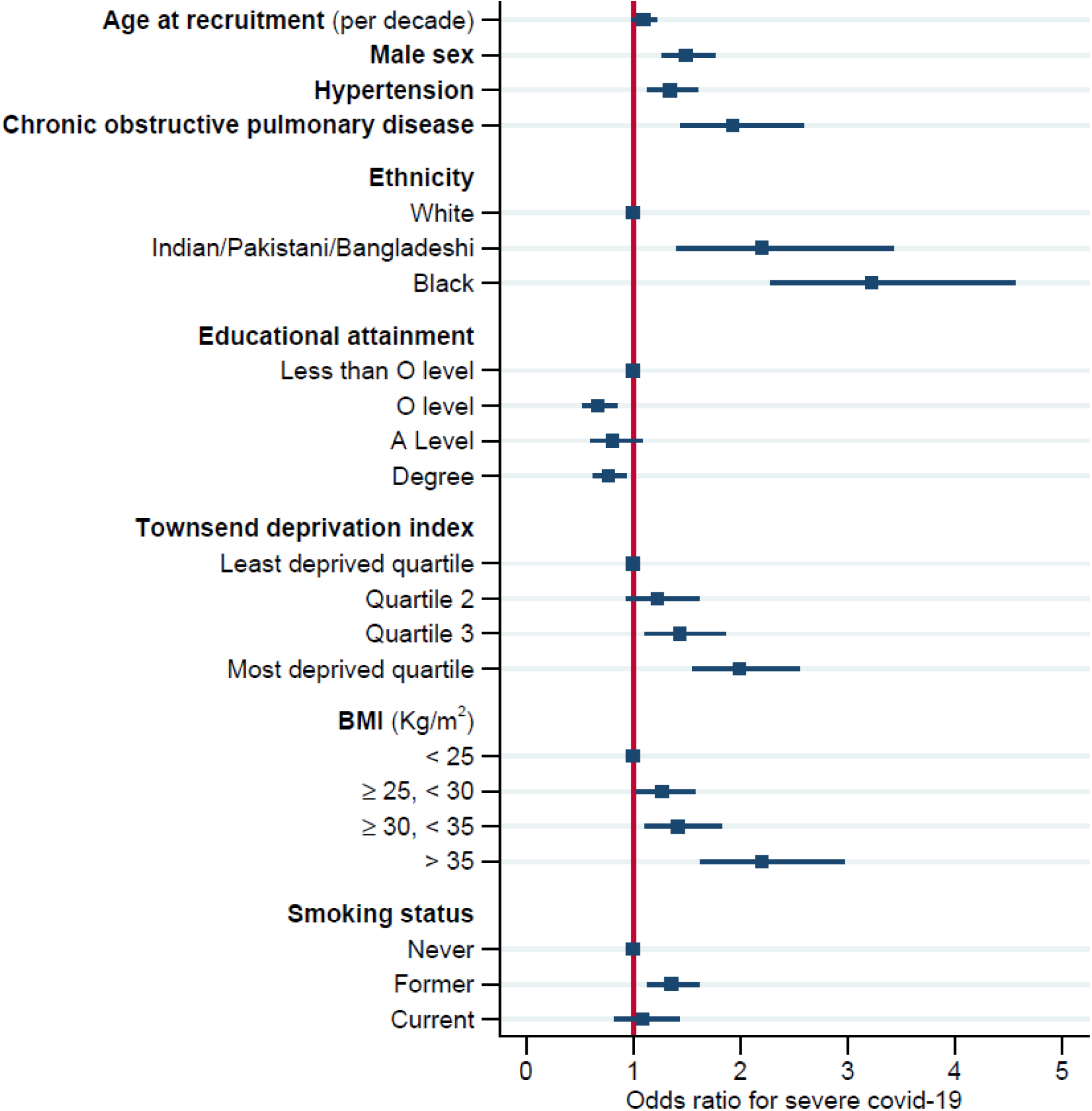
Forest plot for the parsimonious multivariable model examining associations with covid-19 hospitalisation (n=406,793). Numeric values for the coefficients and P-values are presented in supplementary table S4.

## Discussion

Our study is one of the first examining risk factors for severe covid-19 prospectively and at a population-level. The major independent risk factors were male sex, non-White ethnicity, residence in an area with high socioeconomic deprivation, no educational qualifications, smoking status, high BMI, and history of hypertension or COPD. Some of the associations are similar to those reported in hospital case series,^4,15–17^ lending support to the validity of the data linkage and assumptions made in our analyses. However, population-based comparisons enabled estimates of the magnitude of the individual risk factors and, in addition, assessments of their independent relationship with risk of covid-19 infection in multivariable models.

The increased risk for men compared to women is notable and well documented in hospital case series from China,^15^ Europe^18^ and the US.^4,16^ While this has been partly attributed to men having a higher prevalence of risk factors such as smoking, hypertension, diabetes, COPD and obesity, in the current multivariable analyses, male sex was associated with increased risk independently of all these factors. Reports indicate that males show a higher susceptibility to many infectious disease, including some limited evidence in humans for SARS^19^ and MERS,^20^ as well as animal studies.^21^ Differential vulnerability between males and females may come from differences in exposure and social habits, immune responses,^22^ hormonal modulation,^23^ or a combination of these factors.

The most striking and strongest risk factor for severe covid-19 infection we observed was ethnicity. Ethnic differences in covid-19 infection are well documented. A greater susceptibility to severe disease and mortality in non-White ethnic groups has been recorded in both the US^8,9,24^ and the UK.^3,5,7^ We observed over two-fold increased risk of severe covid-19 in Indian/Pakistani/Bangladeshi ethnic groups and over three-fold increased risk in Black ethnic groups. Further analyses stratifying Blacks into African and Caribbean subgroups yielded similar effects for both groups (OR 2.95 and 3.38 respectively, both *P*<0.001). This suggests the increased risk is evident for both groups, contrary to a report that the association may be mainly driven by Caribbeans.^6^ Suggested explanations for these differences have included the higher prevalence of risk factors including diabetes, hypertension and obesity in Black and Asian people, and differing socioeconomic factors. Nonetheless, we still observed an increased risk after adjusting for all these risk factors. While we were only able to examine risk of hospitalised severe covid-19 infection, and not mortality, these observations reinforce the urgent need to understand the reasons for this marked ethnic difference.

The independent association with residential socioeconomic deprivation was notable. There was a marked gradient of increasing risk with increasing deprivation and 40% of cases were in the most deprived quartile. The strength of this finding is particularly surprising given that the score is based on residential postcode of the individual, so has substantial measurement error. Similarly for another socioeconomic index, those with no educational qualification had increased risk compared to those with any qualifications. Possible explanations for why people with poorer socioeconomic indices might be more susceptible to severe covid-19 include that they are a surrogate measure for poorer housing and overcrowding, or need to use public transport; the higher risk of urban compared to rural communities has been reported, although many rural areas are also highly deprived on this index. Another possibility is poor overall general resilience relating to poorer nutrition.

As with the reports from case series,^17^ we observed that a history of hypertension was a strong risk factor for severe covid-19. There has been much debate as to reasons for this, including the hypothesis that human pathogenic coronaviruses bind to their target cells through angiotensin-converting enzyme 2 (ACE2) which is expressed by various epithelial cells. It is suggested that individuals who are treated with ACE inhibitors and angiotensin II type-1 receptor blockers (ARBs), such as those with hypertension, have increased expression of ACE2 which may increase the risk of developing severe and fatal covid-19.^25^ In this cohort, we were able to explore in detail the use of seven different classes of antihypertension medication. Though in univariable analyses all classes of antihypertensive medication were associated with increased risk, in detailed multivariable analyses none of these were independently associated with severe covid-19 infection after adjusting for hypertension history, age, sex and ethnicity; rather it was the number of antihypertensive medications in use that was significantly related, which is probably a surrogate for severity of hypertension. This suggests it is more likely that hypertension per se, in particular hypertension severe enough to require multiple medications, that is the main risk factor for severe covid-19. These results lend some reassurance to the consensus reports and reviews^12,26,27^ that there is no strong evidence to implicate ACE inhibitors or ARB use in covid-19 severity and that individuals should continue with such medication to control hypertension. There were no strong associations of systolic or diastolic blood pressure with covid-19, suggesting that a history of hypertension or use of antihypertensives are more robust indicators of risk than one-off blood pressure measures. Nevertheless, questions remain as to why hypertension should be such a strong risk factor for covid-19.

In marked contrast, while diabetes was crudely associated with severe covid-19, there was no significant association after adjusting for other comorbidities; it is possible the crude association is confounded by hypertension or higher BMI in these individuals. The only other comorbidity independently related to severe covid-19 was a history of COPD. Interestingly, we did not find an independent association with asthma; this is of interest given its prevalence.

Some hospital data suggests that obese individuals have a worse outcome after admission with covid-19.^28^ Our data suggest that more obese people are also at higher chance of developing disease requiring hospitalisation in the first place. BMI measured 12 years previously was a strong risk factor for hospitalisation with covid-19; there was a marked dose response relationship with a two-fold increased risk in those with BMI >30 kg/m^2^ and more than three-fold risk in those with BMI >35 kg/m^2^ compared to those with BMI <25 kg/m^2^. Several mechanisms for obesity-mediated risk have been suggested, including higher propensity to thrombosis, poorer immune response or excess inflammatory response.^29^ Obesity decreases key operating lung volumes and this effect appears particularly pronounced in men.^30^

We observed a strong association with increased risk of severe covid-19 in ever smokers, indicating a harmful impact of smoking on the risk of developing severe covid-19. This risk was mainly observed in the former rather than current smokers; this could be related to the low prevalence of current smokers in this cohort, or that participants still smoking represent a healthier subset of all prior smokers in that some former smokers may have given up due to poorer health.

Surprisingly, age was not significant in the final multivariable model. It could be that association with age is largely driven by comorbidities. While older age is a strong risk factor for covid-19 mortality, age in UK Biobank (which has a restricted age range) may not be a risk factor for acquiring covid-19. Another possible explanation would be the admission policies for hospitals in UK; the very oldest individuals might not be admitted to hospital.

### Limitations

A major issue is the classification of cases and controls. The ascertainment of severe covid-19 cases in the UK Biobank cohort was almost certainly an underascertainment. We identified cases through the national Public Health database which collects data for all covid-19 tests, both positive and negative. After the initial contact tracing policy, from 16^th^ March 2020, testing was reserved for individuals hospitalised with covid-19. This was due to limited test availability and to protect services from being overwhelmed by people with mild disease. Individuals identified with positive covid-19 tests from the UK Biobank cohort therefore represent the hospitalised and most severe cases. There will be individuals in the comparison control population who are susceptible to severe covid-19 infection but who have been uninfected to date. These numbers are likely to be relatively small compared to the overall denominator. In any case, misclassification of controls is likely only to attenuate any associations observed and would not explain our significant findings. Furthermore, our results are in keeping with hospital-based cohort reports. While this is one of the first studies to prospectively examine community-level associations with severe infection, we did not have data to assess subsequent mortality; future linkage to hospital outcome data will enable these analyses. While the diversity of the UK population permits adequately powered analyses of ethnicity, our findings may not apply to non-UK populations.

The major strengths of this study are the large UK population-based cohort, with the ability to quantify and assess risks of severe covid-19 infection in cases and controls in relation to characteristics measured years prior to infection in 2020. In addition, many risk factors for covid-19 documented in the literature are highly correlated and it is not clear which may be independently related to risk. The large numbers of covariables available in this cohort also enabled multivariable adjustment, permitting assessment of independent risk factors. In particular, the marked increased susceptibility of non-White ethnic groups including Black and Indian/Pakistani/ Bangladeshi, men, and those with a history of hypertension or obesity, as well as the increased risks observed with socioeconomic residential deprivation and low education, require further investigation. Understanding why these factors confer increased risk of severe covid-19 in the population may help elucidate the underling mechanisms as well as inform policy to prevent this disease and its consequences.

## Data Availability

Researchers wishing to access UK Biobank data can register and apply at https://www.ukbiobank.ac.uk/

## Acknowledgements

This research has been conducted using the UK Biobank Resource under Application Number 36741.

